# A Novel Transfer Learning Model for Predictive Analytics using Incomplete Multimodality Data

**DOI:** 10.1101/2020.04.23.20077412

**Authors:** Xiaonan Liu, Kewei Chen, Teresa Wu, David Weidman, Fleming Y. M. Lure, Jing Li, The Alzheimer’s Disease Neuroimaging Initiative (ADNI)

## Abstract

Multimodality datasets are becoming increasingly common in various domains to provide complementary information for predictive analytics. In health care, diagnostic imaging of different kinds contains complementary information about an organ of interest, which allows for building a predictive model to accurately detect a certain disease. In manufacturing, multi-sensory datasets contain complementary information about the process and product, allowing for more accurate quality assessment. One significant challenge in fusing multimodality data for predictive analytics is that the multiple modalities are not universally available for all samples due to cost and accessibility constraints. This results in a unique data structure called Incomplete Multimodality Dataset (IMD) for which existing statistical models fall short. We propose a novel Incomplete-Multimodality Transfer Learning (IMTL) model that builds a predictive model for each sub-cohort of samples with the same missing modality pattern, and meanwhile couples the model estimation processes for different sub-cohorts to allow for transfer learning. We develop an Expectation-Maximization (EM) algorithm to estimate the parameters of IMTL and further extend it to a collaborative learning paradigm that is specifically valuable for patient privacy preservation in health care applications of the IMTL. We prove two advantageous properties of IMTL: the ability for out-of-sample prediction and a theoretical guarantee for a larger Fisher information compared with models without transfer learning. IMTL is applied to diagnosis and prognosis of the Alzheimer’s Disease (AD) at an early stage of the disease called Mild Cognitive Impairment (MCI) using incomplete multimodality imaging data. IMTL achieves higher accuracy than competing methods without transfer learning.

## 1. Introduction

Multimodality datasets are becoming increasingly common in various domains to provide complementary information for predictive analytics. For example, in health care, images of different types such as structural magnetic resonance imaging (MRI) and fludeoxyglucose positron emission tomography (FDG-PET) provide complementary information about the organ of interest, which allows for building a predictive model to accurately diagnosing a certain disease (Jack et al. 2009; Lowe et al. 2009; Clark et al. 2011). In manufacturing, data collected from multiple different types of sensors provide complementary information about the process and product, allowing for more accurate assessment of process and product quality (Basir and Yuan 2007).

One important challenge for integration of multimodality datasets in building a predictive model is that the multiple different modalities are not universally available for all the samples. Take the diagnosis of the Alzheimer’s disease (AD) – a fatal neurological disorder – using multimodality images as an example. Figure 1 shows the special “incomplete multimodality dataset (IMD)” we are focusing on in this paper, which includes three complementary image modalities, i.e., MRI, FDG-PET, and amyloid-PET for detection of AD at an early stage of the disease called Mild Cognitive Impairment (MCI) (Jack et al. 2012). In the recently published expert consensus criteria by the National Institute of Aging and Alzheimer’s Association, the use of multimodality images for early detection of AD has been highly recommended (Albert et al. 2011). In Figure 1, each sub-cohort consists of patients who have the same availability of modalities. Different sub-cohorts have different missing modality patterns. The reasons for the existence of IMD are multifold: some imaging equipment such as PET is costly and only available in limited clinics; some modalities are not accessible to patients due to insurance coverage; it is not safe to put patients with some pre-existing conditions through a certain imaging examination.

**Figure 1.**
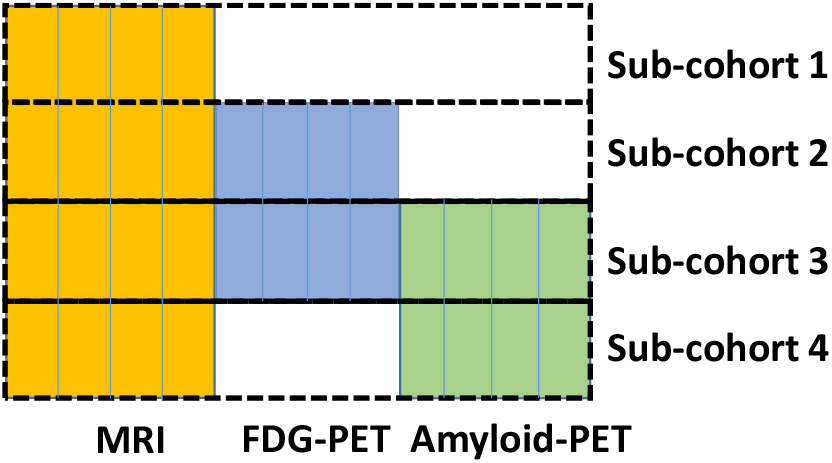
An example of the incomplete multimodality dataset (IMD), in which MRI, FDG-PET, and amyloid-PET are considered as three modalities. Columns within each modality represent features extracted from the image. Each sub-cohort consists of patients with the same availability of modalities.

If we applied existing methods to model IMD, there would be three options.

1. Filling in missing data using some imputation algorithms (He et al. 2017; Ordóñez Galán et al. 2017). Because an IMD dataset misses the entire modality/modalities not individual features within a modality, there are too many missing values to fill in. The resulting dataset may have poor quality and thus negatively affecting performance of the subsequent predictive model based on this dataset.
2. Separate modeling (SM). Since each sub-cohort has different availability for the modalities, SM builds a separate predictive model for each sub-cohort using the available modality/modalities within that sub-cohort. The limitation of SM is obvious: because each model can only use the data specific to the corresponding sub-cohort, sample size shortage may prevent building a robust model.
3. All available data modeling (AADM). To build a model for each sub-cohort *l*, one can incorporate data from another sub-cohort *l′* whose available modalities include those in *l*. For example, to build a model for sub-cohort 2, one can combine the data of MRI and PDG-PET in sub-cohort 3. Because all the available data is used regardless of which sub-cohort the data resides in, this approach is called AADM. Compared with SM, AADM alleviates the sample size shortage. However, it requires data pooling from different sub-cohorts. In reality, different sub-cohorts likely correspond to different health institutions or hospitals, data pooling is not easy due to the concern of patient privacy. Furthermore, it is known that AADM may not produce statistical consistent estimators; when it is used to estimate a covariance matrix, the estimate may not be positive definite (Little and Rubin 2002).

In this paper, we propose a novel Incomplete-Multimodality Transfer Learning (IMTL) model to tackle the limitations of existing methods. IMTL models all the sub-cohorts simultaneously under a unified framework. In this way, knowledge obtained from the modeling of each sub-cohort can be “transferred” to help the modeling of other sub-cohorts. This makes IMTL a transfer learning model. Compared with SM, IMTL is not limited by the sample size of each sub-cohort. Compared with AADM, IMTL estimates model parameters in an integrated manner, which overcomes the limitations of AADM in the lack of positive definiteness and consistency guarantees. Furthermore, we propose two algorithms for parameter estimation of IMTL: with and without data pooling. The latter is a computational framework that includes iterative communication between a global learner and local learners residing within each sub-cohort/institution. This allows for between-institutional collaborative model estimation without the need for data pooling. This is particularly important for patient privacy preservation in health care applications of IMTL. Finally, we would like to stress that although IMTL is developed in the context of multimodality data in health care, it can be effortlessly extended to other non-medical domains that fusion of multimodality datasets is common and much needed, including but not limited to manufacturing (Basir and Yuan 2007) and transportation (Xia, Li, and Shan 2013).

The remainder of the paper is organized as follows: Sec. 2 provides a literature review. Sec. 3 presents the development of IMTL. Sec. 4 investigates unique properties of IMTL. Sec. 5 presents case studies. Sec. 6 is the conclusion.

## 2. Literature review

This paper primarily intersects with the research area of statistical and machine learning models using data missed in chunks of modalities, termed as IMD in this paper. To our best knowledge, this area only has limited work. In what follows, we review each related paper in detail.

Yuan et al. (2012) proposed an incomplete multisource feature learning method (iMSF), which used an *l*_21_ penalty to enforce same features within each modality to be selected across different sub-cohorts. One limitation of iMSF is that it cannot do “out-of-sample prediction”. That is, if a modality-wise missing pattern is not included in training data, iMSF cannot make a prediction on new samples with that missing pattern. Also, the *l*_21_ enabled feature selection scheme is most effective if different modalities have little correlation. To overcome the limitations of iMSF, Xiang et al. (2014) proposed an incomplete source-feature selection (ISFS) model. The main idea was to estimate a set of common coefficients across different sub-cohorts and specific coefficients to account for the uniqueness of each sub-cohort. To gain this flexibility, ISFS needs to estimate many parameters.

Thung et al. (2014) developed a matrix completion method, which selected samples and features in the original dataset to produce a smaller dataset. This was done by using the group-lasso based multitask learning algorithm twice on features and samples, respectively. Then, standard missing data imputation algorithms were applied to the reduced dataset and classifiers were built on the imputed data. While the proposed idea of data reduction is novel, imputation would still have to be used.

Liu et al. (2017) proposed a view-aligned hypergraph learning (VAHL) method. VAHL divided the dataset into several views according to the availability of different modalities. A hypergraph of subjects was constructed on each view. Then, the hypergraphs were fused by a view-aligned regularizer under a classification framework. VAHL had a novel perspective of exploiting subject relationship using hypergraphs to naturally get around the issue of missing modalities. Also because of this “subject” perspective, the model has to be re-trained from scratch every time new data are becoming available. Also, VAHL has many parameters to estimate.

Li et al. (2014) proposed a deep learning (DL) framework specifically for imaging data. The basic idea was to train a 3-D convolutional neutral network (CNN) to establish a voxel-wise mapping from an MRI image to an FDG-PET image based on a dataset with both images available. Then, the CNN could be used to create a “pseudo” FDG-PET from an MRI image for any patient whose FDG-PET is missing. To perform diagnosis or prognosis for a patient, both MRI and FDG-PET (real or pseudo) would be used. This work represents one of the pioneers that introduced DL into imaging-based AD research. On the other hand, because MRI measures brain structure and FDG-PET measures brain function, crafting one from the other may not be biologically valid even though this is possible from a pure data-driven perspective. Further, there is a concern of uncertainty propagation as the uncertainty in establishing the voxel-wise mapping between MRI and FDG-PET will propagate to the uncertainty of the pseudo FDG-PET, which further affects the diagnosis and prognosis based on the pseudo FDG-PET. Also, this approach was developed to model two image modalities and is not directly applicable to datasets with more than two modalities and complicated missing patterns (e.g., Figure 1).

In summary, limited work has been done to develop statistical models for IMD data. All the above-reviewed models, despite their specific weakness, share some common limitations: 1) Most models cannot do out-of-sample prediction, which limits broader utilization; 2) Model estimation needs data pooling from different sub-cohorts. If the sub-cohorts correspond to different health institutions, which is typically the case, protection of patient privacy is a concern. Also, the institutions have to establish data sharing agreement before the modeling can take place, which is a lengthy process if not impossible. 3) While showing empirically good performance on specific datasets, there is a lack of theoretical study on why the performance is guaranteed.

## 3. Development of the Incomplete-Multimodality Transfer Learning (IMTL) model

For notational simplicity, we present our model development in the context of three modalities, while the model is generalizable to other numbers of modalities. For example, the three modalities can be MRI, FDG-PET and amyloid-PET as shown in Figure 1. Note that in Figure 1, we assume that MRI is available to patients in all the sub-cohorts. This is a valid assumption because MRI is in the standard care of AD. Under this structure, there are four patient sub-cohorts corresponding to different availabilities of the modalities: MRI alone; MRI & FDG-PET; MRI & amyloid-PET; all three modalities.

Let *k* be the index for modalities, *k* = 1,2,3; *l* be the index for sub-cohorts, *l* = 1,2,3,4; and *i* be the index for samples/patients, *i* = 1, …, *n*. Denote the sample size of each sub-cohort by 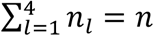. Furthermore, let 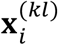 contain features in modality *k* for patient *i* in sub-cohort *l*. Let 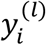 be the response variable for patient *i* in sub-cohort *l*. We propose two IMTL models, one for a continuous response (i.e., a predictive model) and the other for a binary response (i.e., a classification model). Both models are useful in disease diagnosis and prognosis. For example, in diagnosis, 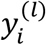 can be a binary variable indicating existence of the disease or a continuous variable representing disease severity. In prognosis, 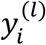 can be a binary variable indicating death or progression to a more advanced stage by a pre-specified future time point or a continuous variable representing the severity the disease will advance into at a future time point or time to death/progression.

### 3.1 IMTL predictive model

#### 1) Formulation and estimation

Consider the joint distribution of 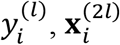, and 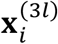 given 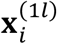 to be multivariate normal, i.e.,

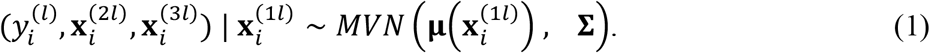

Here, we consider 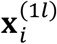 (e.g., features of MRI) to be fixed covariates instead of random variables based on the aforementioned assumption that MRI is in the standard clinical care of AD and thus available to all the patients. While 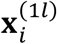 could be considered as random in the most general formulation, doing so would need a joint distribution of 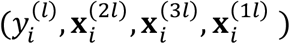, which requires more parameters to be estimated than the proposed formulation in (1), such as the mean vector and variance-covariance matrix of 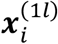 as well as the covariances between 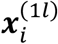 and 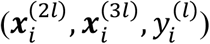.

In (1), **μ**(·) is a vector function of covariates. Although **μ**(·) can take any form in theory, we focus on a linear function in this paper, i.e.,

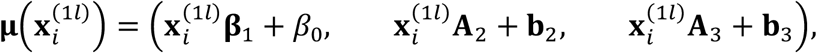

where **β**_1_, *β*_0_, **A**_2_, **b**_2_, **A**_3_, **b**_3_ are coefficients. The covariance matrix **Σ** in (1) can be written in a more explicit format to include sub-matrices of covariances between the response and each modality and between the modalities, i.e.,

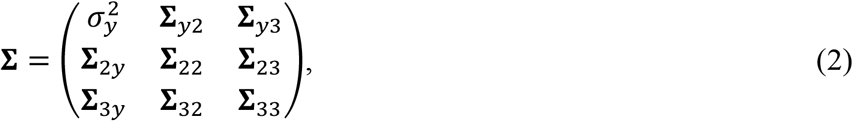

Let **Θ** = (**Σ**, **β**_1_, *β*_0_, **A**_2_, **b**_2_, **A**_3_, **b**_3_) contain all the unknown parameters for the model in (1). Furthermore, let D^*mis*^ and D^*obs*^ contain the missing and observed data corresponding to the data structure in Figure 1, respectively. That is, 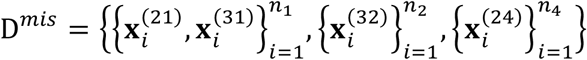 and 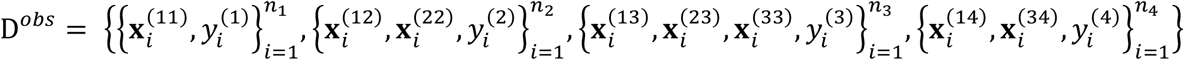. Then, we can write down the complete-data log-likelihood function, i.e.,

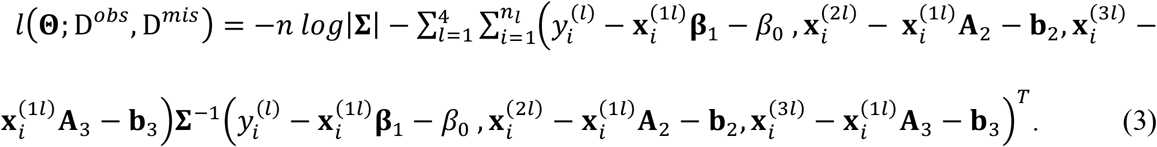

Since *l*(**Θ**; D^*obs*^, D^*mis*^) includes missing data, we resort to the Expectation-Maximization (EM) algorithm. The general EM framework includes an E-step and an M-step. The E-step is to find the expectation of the complete-data log-likelihood function with respect to the missing data given the observed data and the current parameter estimates. In our case, the E-step is to find

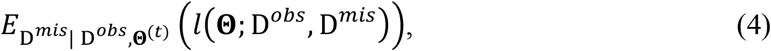

where **Θ**^(*t*)^ contains the parameter estimates obtained at the *t*-th iteration. Then, the M-step is to update the parameter estimates by maximizing the expectation in the E-step, i.e.,

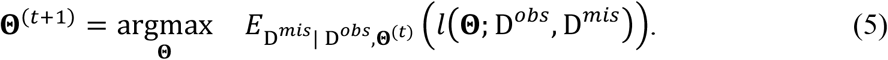

The two steps are iterated until convergence. The challenges in using the general EM framework are to derive the expectation and solve the maximization for a specific model (e.g., IMTL in our case). In what follows, we will develop the details of the E-step and M-step for IMTL.

##### E-step

When the likelihood function is based on a distribution in the exponential family, Little and Rubin (2002) showed that the E-step becomes finding expectations of sufficient statistics. Our likelihood function is based on a multivariate normal distribution in (1). Therefore, the goal of our E-step is to find the sufficient statistics associate with (1) and derive their expectations. Let *S* be a collection of the sufficient statistics. We find that *S* includes the following elements:

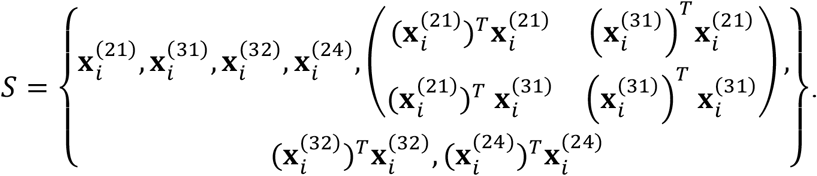

Furthermore, we need to derive the expectation of each element contained in *S*. For example, focus on 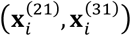 first. We can derive that

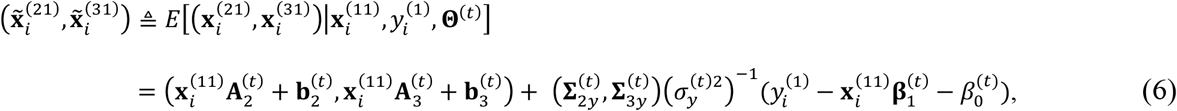

Here, we only show the result and have to skip the derivation process due to space limit. Similarly, the expectations of 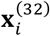 and 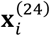 can be obtained as follows:

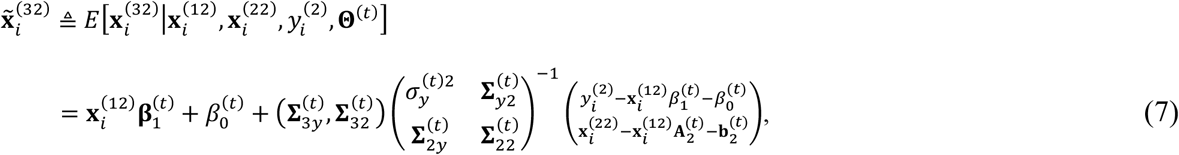

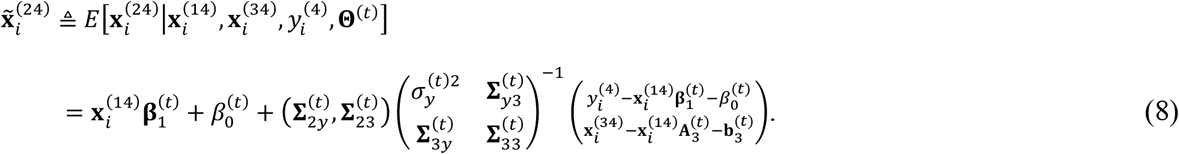

Using (6) -(8), we can further derive the expectations of the 2^nd^-order elements in *S* as:

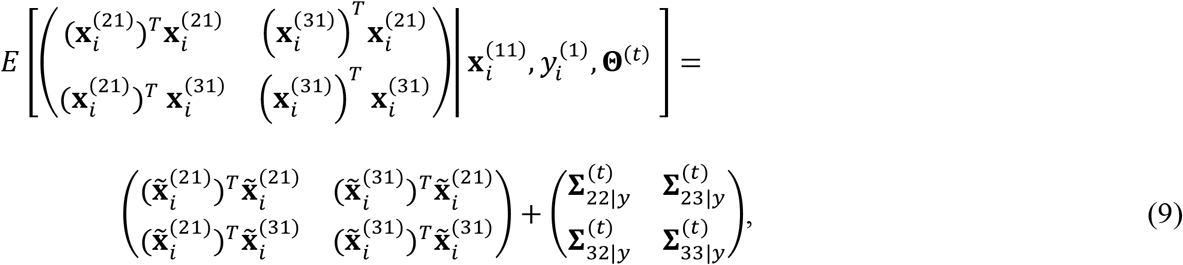

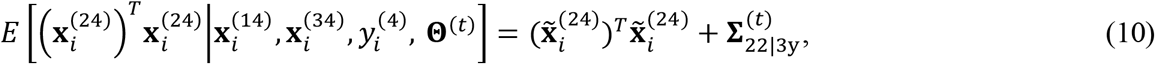

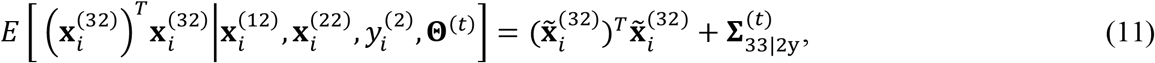

where

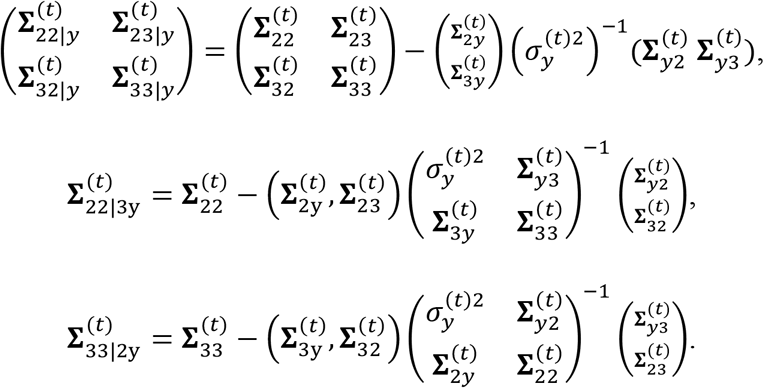

Next, we plug the derived expectations of the sufficient statistics, i.e., (6)-(11), into the expected complete-data log-likelihood function in (4). Through some algebra, (4) can be written as

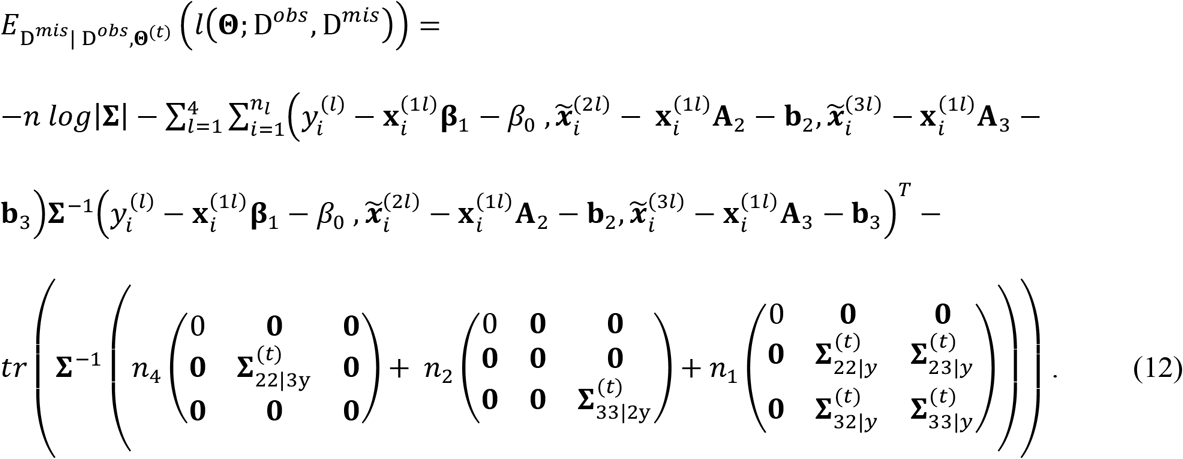

In (12), a notational trick is used, i.e., 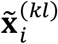 is used to represent the data 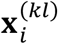 no matter if the data is observed or missing, *k* = 2,3; *l* = 1, …, 4. When the data is observed, e.g., 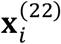, we make 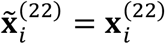. When the data is missing, e.g., 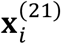, the expression of 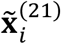 is given in (6). The reason for using this notational trick is to facilitate the maximization in the M-step, which will become apparent in the following discussion.

##### M-step

Split the parameter set **Θ** into two subsets: (**β**_1_, *β*_0_, **A**_2_, **b**_2_, **A**_3_, **b**_3_) and **Σ**. The maximization problem can be solved by taking the partial derivative of the expectation in (12) with respect to each subset and equating the partial derivative to zero, i.e.,

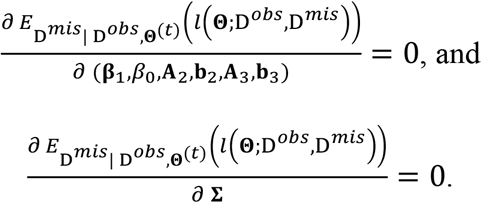

Instead of directly solving these equations, which is computationally involved, we take an indirect approach by first obtaining the least square estimators for the coefficients in the following three regressions:

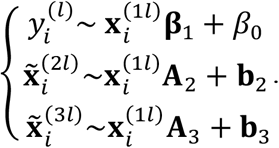

The least square estimators are:

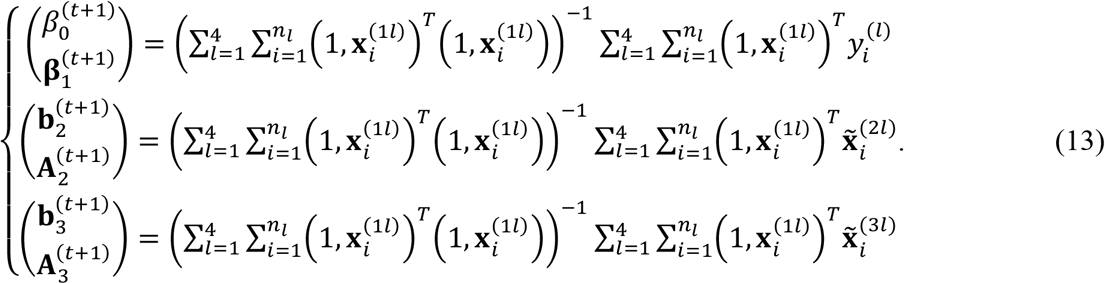

It is not hard to show that these estimators are equivalent to the optimal solutions for (**β**_1_, *β*_0_, **A**_2_, **b**_2_, **A**_3_, **b**_3_) in the M-step. Furthermore, let 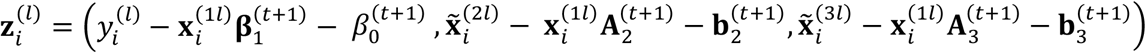. Then, we can obtain the optimal solution for **Σ** as

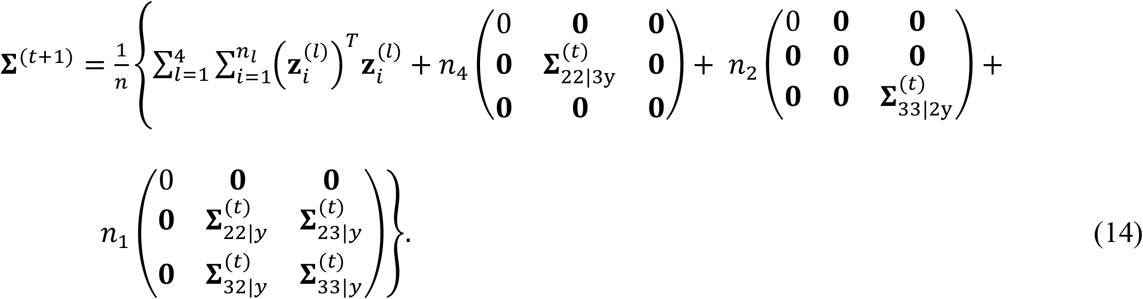

#### 2) Prediction

At the convergence of the above EM iterations, we can obtain the estimated parameters 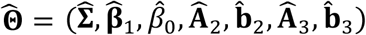. Then, these parameters can be used to predict on new samples. Consider a new sample *i*^∗^. Depending on what available modality/modalities this sample has, we can use the following model to predict the response variable of the sample:

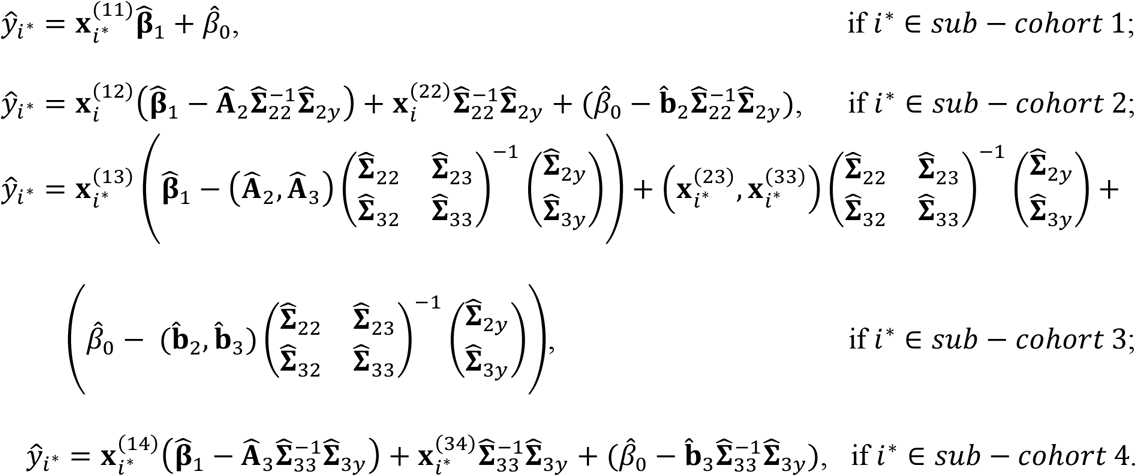

### 3.2. IMTL classification model

In a classification model, 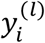 can take the values of 0 or 1 that represent two classes. Within each class, consider the joint distribution 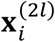 and 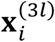 given 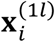 to be multivariate normal, i.e.,

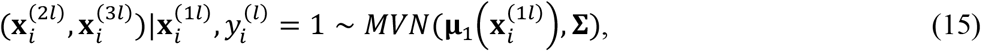

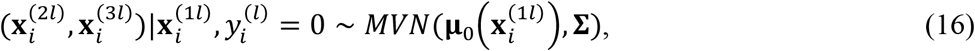

where the class-specific means are linear functions of 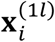, i.e.,

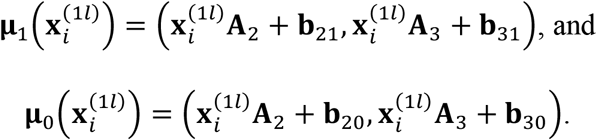

The same covariance matrix, 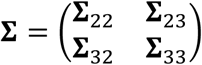, is assumed for the two classes. Furthermore, we consider the distribution of 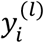 given 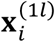 to be Bernoulli, i.e.,

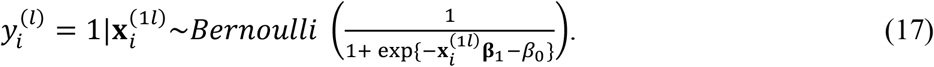

Let 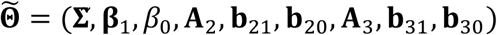 contain the unknown parameters for the model in (15)-(17). We can write down the complete-data log-likelihood function, i.e.,

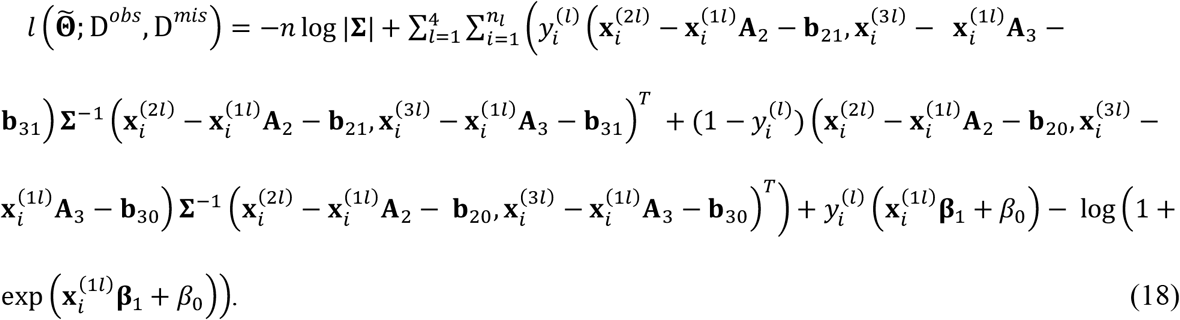

Equation (18) can be decomposed into a logistic regression and a conditional multivariate normal distribution. As a result, we can estimate (**β**_1_, *β*_0_) and the remaining parameters in 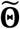 separately. Specifically, (**β**_1_, *β*_0_) are coefficients of the logistic regression model:

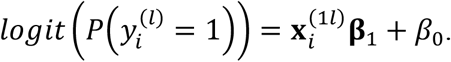

This model does not involve missing data, which means that (**β**_1_, *β*_0_) can be estimated by iteratively reweighted least squares (IRLS) estimation.

Furthermore, let Θ be the parameters in 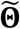 excluding (**β**_1_, *β*_0_). Θ can be estimated by a similar EM algorithm to the predictive model in Sec. 3.1. Please see Appendix B for the formula in the EM algorithm and in the classification models on new samples.

### 3.3. Collaborative model estimation without data pooling

One reason leading to generation of IMD data in health care applications is that each sub-cohort corresponds to a different institution. The availability of modalities varies across the different institutions due to accessibility and cost. In the IMTL models proposed in Sec. 3.1-3.2, model estimation is assumed to happen at a centralized place into which the data from different institutions (i.e., sub-cohorts) have been deposited. This requires a multi-institutional data sharing agreement – a process known to be time- and effort-intensive. A more commonly encountered scenario is that different institutions would like to collaborate on model estimation without having to share their respective patients’ data. In this section, we address the latter scenario by proposing a computational framework for model estimation. This framework uses the same equations as those in Sec. 3.1-3.2, but it computes the equations at the E-step locally at each institution because each equation only involves the data from a single institution. This is shown as the four vertical rectangular boxes in Figure 2. For example, equation (6) in the E-step computes the expectations of two missing modalities in sub-cohort 1, 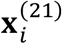 and 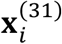, which only involve the data from sub-cohort 1, 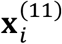 and 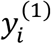. This nice “local” property also holds for other equations in the E-step. At the M-step, the proposed framework combines the locally computed results in a centralized place, which is shown as the horizontal rectangular box at the top of Figure 2. Because these results do not reveal the raw data in each institution, patient privacy is preserved within each institution. The key idea of this computational framework is to consider the M- and E-steps as a global and a local learner, respectively. The global learner resides in a centralized place while the local learners reside in each sub-cohort. A local learner can only “see” the data within the respective sub-cohort and performs computation locally. Results from the local computation, not the data, are sent to the global learner to be combined.

**Figure 2.**
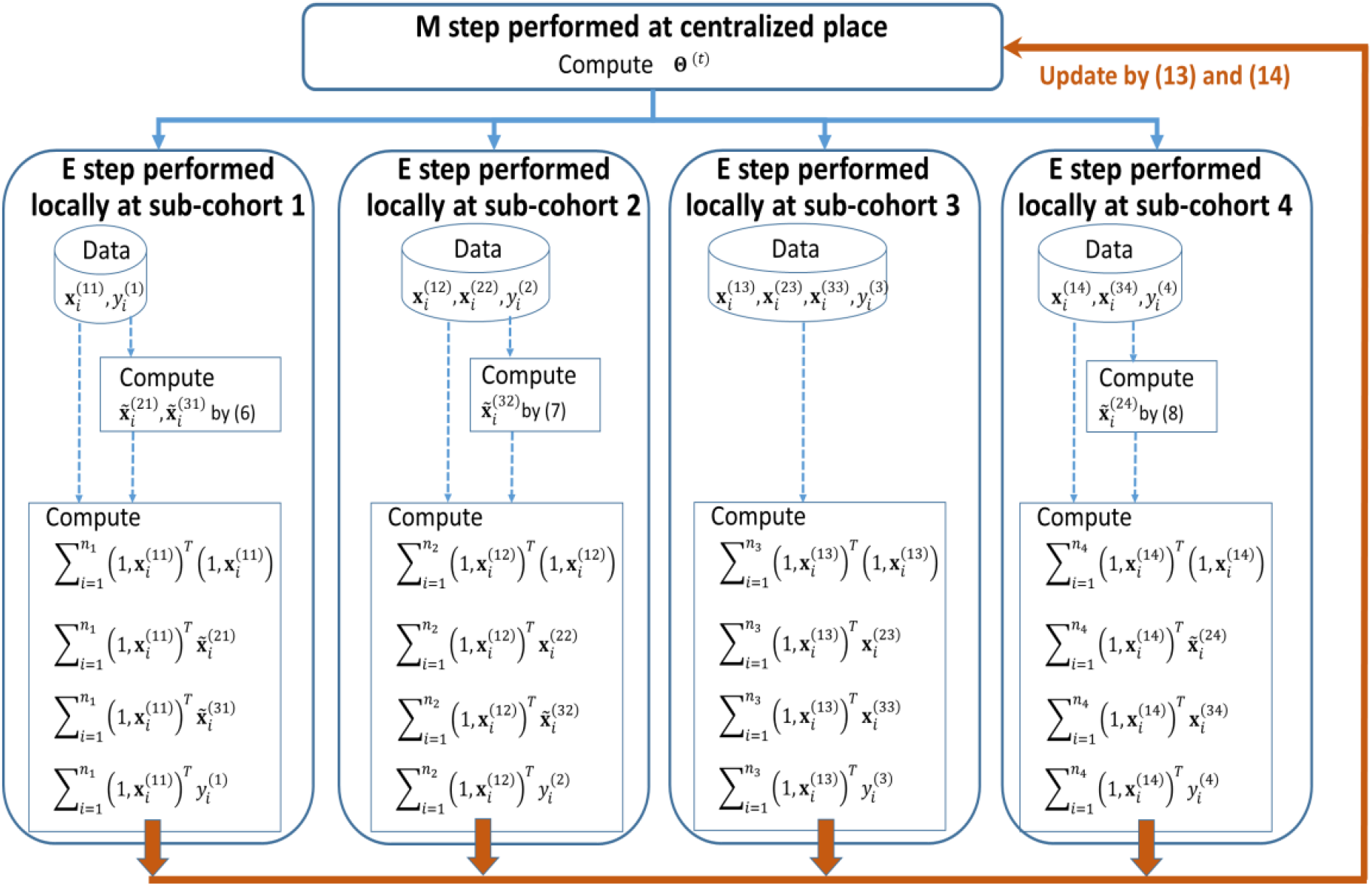
A computational framework for collaborative model estimation of IMTL without data pooling from different sub-cohorts.

Compared with the centralized model estimation in Sec. 3.1-3.2, this computational framework involves communications between the global and local learners. As a result, there may be loss of efficiency due to limited communication bandwidth. On the other hand, this problem can be potentially mitigated because the computations of local learners can be performed in parallel.

### 3.4. Generalization to M modalities

The presentation of IMTL in Sec. 3.1-3.3 was within the context of three modalities based on the consideration of notational simplicity. In this section, we provide the steps of extending IMTL to the general case of *M* modalities: 1) Given a multimodality dataset from an application, subjects (a.k.a. samples) are grouped into sub-cohorts with each sub-cohort having a different pattern of missing modalities. 2) Depending on the type of the response variable, one can decide if the problem to be tackled is regression or classification. For a regression problem, a multivariate normal distribution can be assumed for the modalities and the response. For a classification problem, a multivariate normal distribution can be assumed for the modalities and a Bernoulli distribution can be assumed for the response. For most applications, there is at least one modality available across all the sub-cohorts. If this is the case, the aforementioned distributions can be modified into conditional distributions given the available modality. Next, one can write down the complete-data log-likelihood function under the distribution assumption. 3) In the E-step of the EM algorithm, the key is to identify the sufficient statistics of the log-likelihood function, which include the missing modalities in each sub-cohort, the quadratic term of each missing modality, and pair-wise products between the missing modalities in that sub-cohort (if there is more than one missing modality). Then, one can derive expectations of the sufficient statistics given the observed data and parameter estimates from the previous iteration. Further, these expected sufficient statistics are plugged into the expected complete-data log-likelihood function, which will be maximized in the M-step. 4) In the M-step, an intuitive but mathematically-involved approach is to equate the 1^st^-order partial derivative of each parameter to zero and solve the parameter-wise simultaneous equations to update the parameter set. Alternative approaches can be developed to solve the maximization problem easier, depending on the form of the log-likelihood function. For example, in the three-modality case, we used a notational trick, which allowed us to convert the maximization into the solving of least square estimations.

#### Computational complexity

The proposed EM algorithm for IMTL estimation has analytical solutions in the E-step and M-step. Therefore, the computational complexity primarily comes from the iterations between the two steps until convergence. The complexity of EM iterations has been well-studied in the literature. Furthermore, within the E-step, since there are no iterations but just arithmetic operations based on derived mathematical formula, the complexity primarily depends on how many expectations to compute. Given *M* modalities, the total number of sub-cohorts with missing modalities is *L* = 2^*M*−1^ − 1. Within each sub-cohort, there are two types of expectations to compute, including the mean vector and variance-covariance matrix of the missing features. Therefore, the complexity of the E-step can be considered as 2(2^*M*−1^ − 1). Within the M-step, the complexity primarily depends on how many parameters to estimate. Suppose each modality has *p* features. The total number of parameters is 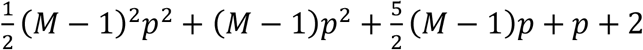.

## 4. Properties of IMTL

In this section, we discuss two unique properties of IMTL: 1) the ability for out-of-sample prediction; 2) a theoretical guarantee for a larger Fisher information compared with models without transfer learning, which explains the superiority of IMTL from a theoretical point of view.

### 4.1. Ability for out-of-sample prediction

**Definition**: Let *D*_*tr*_ denote a training set. Suppose the training samples can be divided into *L* sub-cohorts, where each sub-cohort corresponds to a different missing modality pattern in {𝒫_1_, …, 𝒫_*L*_}. Let *i*^∗^ be a sample in a test set, whose missing modality pattern is 𝒫(*i*^∗^). Assume 𝒫(*i*^∗^) ∉ {𝒫_1_, …, 𝒫_*L*_}. If a model trained on *D*_*tr*_ can be used predict *i*^∗^, the model is called capable of *out-of-sample prediction*.

For example, a training set can include only sub-cohorts 1, 2, and 4 in Fig. 1 while the test set includes sub-cohort 3. It is obvious that the two competing methods to IMTL, i.e., SM and AADM, cannot do out-of-sample prediction. In contrast, IMTL is capable of out-of-sample prediction. Next, we provide an illustrative proof for this capability of IMTL. We focus on the predictive model in Sec. 3.1. Also, for notational simplicity, each modality is assumed to contain one feature.

Consider a sample *i*^∗^ in the test set who belongs to sub-cohort 3. To predict the response variable of this sample, (19) will be used, i.e.,

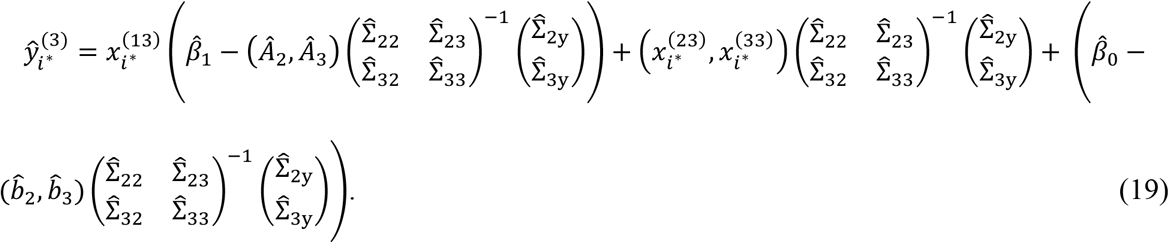

The parameters of the model in (19) are estimated from a training set that includes only sub-cohorts 1, 2, and 4 but not 3. It is easy to understand why estimation of other parameters is possible except Σ_23_. Intuitively, since Σ_23_ is the covariance between features in modalities 2 and 3, one would expect to have at least *some* training data from sub-cohort 3, which have both modalities 2 and 3 available, in order to estimate Σ_23_. However, this is not our case. Therefore, the key to demonstrating that IMTL can do out-of-sample prediction is to demonstrate that the estimation for Σ_23_ is possible by IMTL even without any data from sub-cohort 3 in the training set. To show this, consider the estimation for Σ_23_ by the EM algorithm. At convergence, it can be derived that Σ_23_ can be estimated by (20). The detailed derivation is skipped due to space limit.

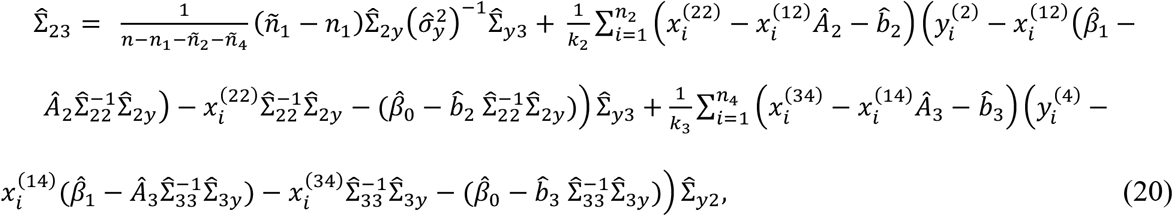

where

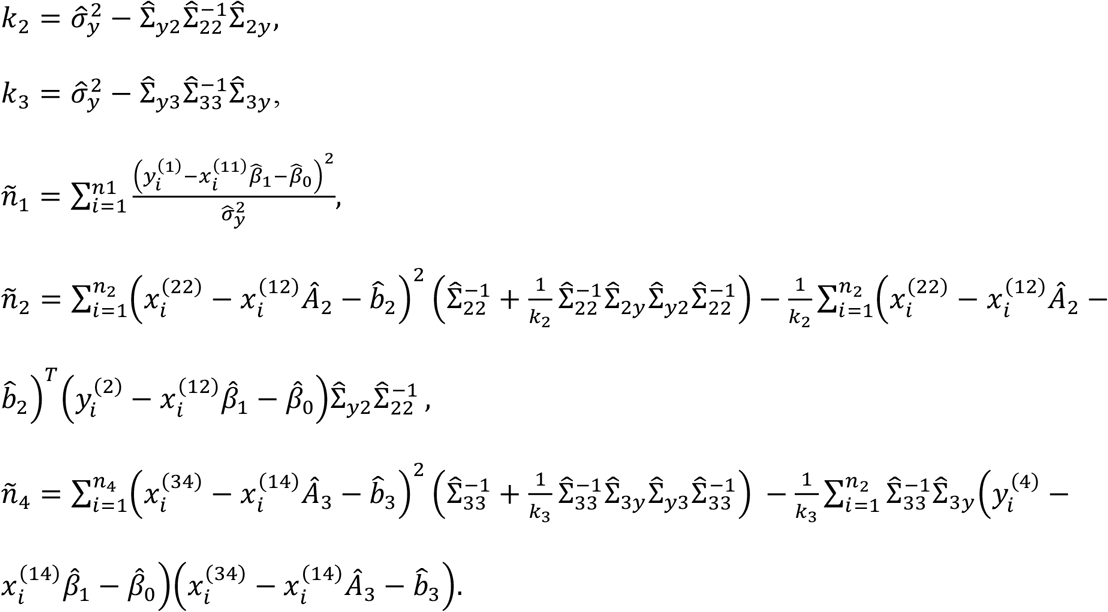

Equation (20) indicates that although training data from sub-cohort 3 are not available, Σ_23_ can be estimated *indirectly* through a summation of three parts: The first part, 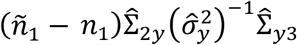, contributes to estimating the covariance between modalities 2 and 3 through exploiting their respective covariances with *y*. The second part leverages the training data in sub-cohort 2, and contributes to estimating Σ_23_ by exploring the covariance between residual modality 2 and residual modality 3. Here, residual modality 2 is modality 2 after factoring out modality 1; residual modality 3 is the residual of the response variable regressing on modalities 1 and 2. Both residual modalities are computed using the training data in sub-cohort 2. Similarly, the third part leverages the training data in sub-cohort 4, and contributes to estimating Σ_23_ by exploring the covariance between residual modality 2 and residual modality 3 that are computed on the training data in sub-cohort 4.

Furthermore, we can explain why estimation of Σ_23_ without sub-cohort 3 is possible from another angle. Using the Law of Total Covariance, Σ_23_ can be decomposed as 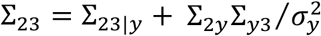. We cannot estimate Σ_23|*y*_ due to the lack of sub-cohort 3. However, Σ_2*y*_, Σ_3*y*_, and 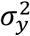 in the second term on the right-hand side can be estimated using the available sub-cohorts. This means that the estimator for Σ_23_ will be biased. While it would be ideal to have data for sub-cohort 3 to mitigate the bias in estimating Σ_23_, our simulation experiments in Sec. 5.1 show that the biased estimator performs well in prediction. In statistical models, biased estimators are used quite often and show good performance in prediction tasks.

### 4.2 Fisher information performance

The next section shows empirical evidence that IMTL outperforms SM and AADM, i.e., models without transfer learning. In this section, we would like to explain the performance improvement from a theoretical standpoint, particularly through comparing the Fisher information of parameter estimates from IMTL and SM/AADM. It is known that Fisher information characterizes the variance of a maximum likelihood estimate, and larger Fisher information means smaller variance. It is our goal to find out if IMTL has larger Fisher information for some parameter estimates than SM/AADM, indicating more robust parameter estimation.

For clarity of presentation, we focus on a two-modality case in deriving the Fisher information for IMTL, SM, and AADM. Modality 1 is available for all patients but modality 2 is missed for some patients. This divides the patients into two sub-cohorts: sub-cohort 1 has modality 1 available but misses modality 2; sub-cohort 2 has both modalities available. Following the notational convention in Section 3, let 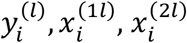 denote the response variable, feature in modality 1, and feature in modality 2 for patient *i* in sub-cohort *l, l* = 1,2. Assume one feature in each modality for notational simplicity. Let D^*mis*^ and D^*obs*^ contain the missing and observed data, i.e.,

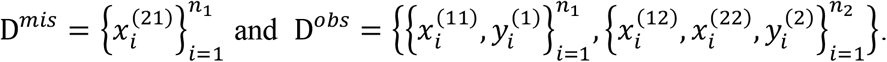

To model this dataset, IMTL assumes a multivariate normal distribution of 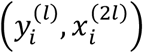 given 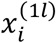, i.e.,

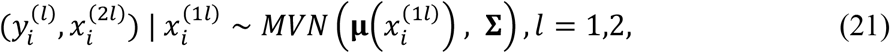

where 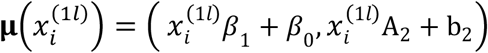 and 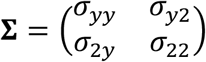. To estimate the parameters of this IMTL model, a similar EM algorithm to that proposed in Section 3.1 can be used. To make predictions on new samples, we can derive the distributions of 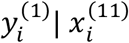 and 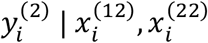 from (21) and use them for predicting samples from sub-cohort 1 and 2, respectively.

If SM is used to model this dataset, there will be two separate models for the two sub-cohorts. Since sub-cohort 2 has no missing modality, the model for sub-cohort 2 has the same form as (21) but with *l* = 2 only. Sub-cohort 1 is separately modeled by a conditional distribution of the response variable given modality 1, i.e.,

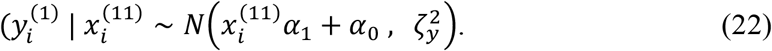

AADM is similar to SM in the sense that two separate models are built for the two sub-cohorts. These models take the same forms as those in SM. However, in estimating the model parameters for sub-cohort 1, AADM uses all available data which includes the data of modality 1 from both sub-cohort 1 and 2, since modality 1 is available for both sub-cohorts. Recall that in SM, only the data from sub-cohort 1 is used. To estimate the parameters of SM/AADM, maximum likelihood estimation can be used since no missing data is involved in the model formulation. To make predictions on new samples, we can use (22) directly if the sample is from sub-cohort 1, and derive and use 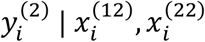 if the sample is from sub-cohort 2.

It can be seen from the above descriptions that IMTL, SM, and AADM share the same model for sub-cohort 2, but they estimate the model parameters in different ways. Theorem 1 compares the Fisher information of the parameter estimates for sub-cohort 2 by the three methods, specifically focusing on the estimates for the elements in the inverse-covariance matrix 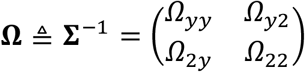

#### Theorem 1

Let *I*_*IMRL*_(*Ω*_22_), *I*_*IMRL*_(*Ω*_2*y*_), *I*_*IMRL*_(*Ω*_*yy*_) be the Fisher information of the estimates for *Ω*_22_, *Ω*_2*y*_, and *Ω*_*yy*_ under IMTL, respectively. Let *I*_*SM*_(∙) and *I*_*AADM*_(∙) be the Fisher information of the estimates for the same parameters under SM and AADM, respectively. Then,

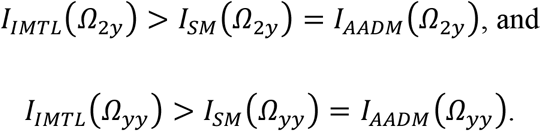

Furthermore,

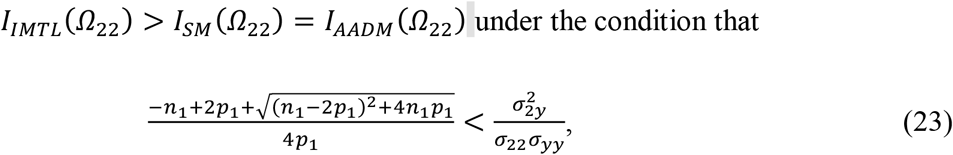

where *n*_1_ is the sample size of sub-cohort 1 and *p*_1_ is number of features in modality 1.

Please see the proof in Appendix A. Theorem 1 shows that the Fisher information for the estimates of Ω_2*y*_ and *Ω*_*yy*_ under IMTL is greater than SM/AADM unconditionally. This relationship holds for the estimate of *Ω*_22_ under the condition given in (22). This condition is worthy of further discussion. Specifically, if considering *n*_1_ and *p*_1_ to be fixed (i.e., the left side of (22) is a constant), Theorem 1 indicates that the correlation between modality 2 and the response variable must be sufficiently large (i.e., larger than the constant) in order for IMTL to have a larger Fisher information for the estimate of *Ω*_22_ than SM/AADM. This means that IMTL will be most effective when the modality with missing data is a significant predictor for the response. If the modality contains largely noise with little predictive value, IMTL may not perform as well as models without transfer learning because it runs the risk of transferring noise and thus hurting the model performance. This problem is known as “negative transfer” in the literature (Pan and Yang 2010).

## 5. Application case study

In this section, we apply IMTL to simulated and real-world datasets. Simulation experiments are presented in Sec. 5.1, with purposes of demonstrating the out-of-sample prediction ability of IMTL, which the competing methods (i.e., SM and AADM) do not possess. Sec. 5.2 presents an application of AD diagnosis and prognosis of MCI patients using the Alzheimer’s Disease Neuroimaging Initiative (ADNI) dataset. Here, “diagnosis” means detection of the existence of AD pathology in the brain of an MCI patient. “Prognosis” means prediction if an MCI patient will progress to AD by a certain year of interest, e.g., 6 years. Both tasks are important for treatment and management of the patients.

### 5.1. Simulation experiments

#### 1) Out-of-sample prediction by IMTL predictive model

We conduct simulation experiments for the IMTL predictive model and classification model. For the predictive model, we first generate data for three modalities, i.e., 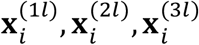, from a zero-mean multivariate normal distribution *MVN*(0, **Σ**). The number of features in each modality is set to be *p*_1_ = 10, *p*_2_ = *p*_3_ = 5, which are close to the size of features in the real-world data presented in Sec. 5.1. All diagonal elements of **Σ** are set to be one. **Σ** includes two parts: within-modality correlation and between-modality correlation. The former has been found to have little impact on the model performance and therefore is set to be 0.6. We investigate two settings for between-modality correlation: 0.6 and 0, which represent moderately strong correlation and no correlation. Furthermore, we investigate two training sample sizes: 300 and 150.

Once the data for features are generated, we generate the response variable *y*^(*l*)^ by a linear model, 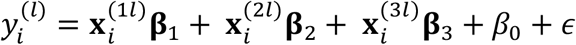. Here, *β*_0_= 2; elements in **β**_1_, **β**_2_, **β**_3_ are set to be 0.2; *ϵ*∼*N*(0,1). Then, the simulated training data are equally separated into three sub-cohorts, *l* = 1, 2, 4, corresponding to sub-cohorts 1, 2, and 4 in Fig. 1. To obtain the incomplete modality pattern in each sub-cohort, we remove the training data of modalities 2 and 3 for sub-cohort 1, remove modality 3 for sub-cohort 2, and remove modality 2 for sub-cohort 4. Because our intention of this experiment is to assess the out-of-sample prediction capability of IMTL, we generate data in a test data that includes only sub-cohort 3, i.e., all modalities are available. The sample size of the test set is 100.

IMTL is trained on the training set that includes only data from sub-cohorts 1, 2, and 4. Then, the trained model is used to predict on the test set that only includes samples from sub-cohort 3. The predicted response variables of the test set is compared with the true responses to compute a prediction mean square error (PMSE) and a Pearson correlation (PC). We repeat the entire experiment for 100 times. Table 1 summarizes the results. As expected, increasing the training sample size significantly improves PMSE and PC (p<0.001). The correlation between modalities also helps improves PMSE and PC (p<0.001). This is consistent with the theoretical discovery in Sec. 4.1, in which we found that the key to out-of-sample prediction was to be able to estimate Σ_23_ from the training data. From (20), it is known that the estimation of Σ_23_ is affected by the correlation between modality 2 and 3. Even though the training data does not include samples with both modality 2 and 3 available, Σ_23_ can still be estimated indirectly by IMTL through exploiting the between-modality correlation and the relationship between modalities and the response variable.

**Table 1A.** Out-of-sample prediction accuracy on the test set with different training sample sizes (between-modality correlation is kept as 0.6 in both settings)

| Training size | PMSE: *ave(std)* | PC: *ave(std)* |
| --- | --- | --- |
| 300 | 1.174 (0.175) | 0.945 (0.010) |
| 150 | 1.469 (0.289) | 0.931 (0.017) |

**Table 1B.** Out-of-sample prediction accuracy on the test set with different between-modality correlations (training sample size is kept as 300 in both settings)

| Between-modality correlation | PMSE: *ave(std)* | PC: *ave(std)* |
| --- | --- | --- |
| 0.6 | 1.174 (0.175) | 0.945 (0.010) |
| 0 | 1.300 (0.187) | 0.866 (0.028) |

#### 2) Out-of-sample prediction by IMTL classification model

The data generation process of this experiment is the same as the previous section except that we use a logistic regression model to link the response variable with predictors/features. Specifically, we first simulate a linear predictor 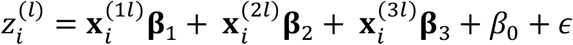. Then, 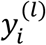 is generated from a Bernoulli distribution with success probability equal to 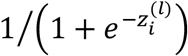. Test accuracy is reported as the Area Under the Curve (AUC). Table 2 summarizes the results. Doubling the training sample size does not seem to dramatically improve the AUC although this improvement is still statistically significant (p<0.001). The correlation between modalities also helps improve the AUC significantly (p<0.001).

**Table 2A.** Out-of-sample classification accuracy on the test set with different training sample sizes (between-modality correlation is kept as 0.6 in both settings)

| Training size | AUC: *ave(std)* |
| --- | --- |
| 300 | 0.882 (0.037) |
| 150 | 0.832 (0.05) |

**Table 2B.** Out-of-sample classification accuracy on the test set with different between-modality correlations (training sample size is kept as 300 in both settings)

| Between-modality correlation | AUC: *ave(std)* |
| --- | --- |
| 0.6 | 0.882 (0.037) |
| 0 | 0.781 (0.046) |

### 5.2. Early diagnosis and prognosis of AD

#### 1) Introduction to ADNI

ADNI (http://adni.loni.ucla.edu) was launched in 2003 by the NIH, FDA, private pharmaceutical companies, and non-profit organizations, as a $60 million, 5-year public-private partnership. The primary goal of ADNI has been to test whether MRI, PET, other biological markers, and clinical and neuropsychological assessment can be combined to measure the progression of MCI and early AD. Determination of sensitive and specific markers of very early AD progression is intended to aid researchers and clinicians to develop new treatments and monitor their effectiveness, as well as lessen the time and cost of clinical trials. The Principal Investigator of this initiative is Michael W.Weiner, MD, VA Medical Center and University of California-San Francisco. ADNI is the result of efforts of many co-investigators from a broad range of academic institutions and private corporations, and subjects have been recruited from over 50 sites across the US and Canada. For up-to-date information, see http://www.adni-info.org/.

#### 2) Patient inclusion and diagnostic/prognostic end points

Our study includes 214 MCI patients from ADNI through our collaborative intuition, Banner Alzheimer’s Institute (BAI), with which two co-authors are affiliated. BAI is a member of ADNI PET core (PI, William Jagust UC Berkeley). Multimodality image data include MRI, FDG-PET, amyloid-PET, which follow the IMD structure in Fig. 1. Each sub-cohort has the same sample size. For *diagnosis*, we use A*β* positivity is an indicator for high-risk AD. We follow the recommendation by Fleisher et al. (2011) and use a threshold of mean SUVR greater than or equal to 1.18 to define A*β* positivity. According to this criterion, there are 87 and 127 patients in class 1 (high-risk) and 0 (otherwise). For *prognosis*, the purpose is to predict when an MCI patient will convert to AD. We searched the ADNI database for the 214 patients from the time when their imaging data were collected up to six years’ follow up, and found that 46 converted to AD, i.e., there are 46 converters (class 1) and 168 non-converters (class 0).

#### 3) Image processing and feature computation

For MRI images, we use the FreeSurfer (http://surfer.nmr.mgh.harvard.edu/) software to extract volumetric measurements for pre-defined regions of interest (ROIs). We focus on three ROIs including hippocampal, ventricle, and entorhinal volumes relative to intracranial volume. All three have been widely reported to be related to AD (Devanand et al. 2007; Thompson et al. 2004). Both FDG-PET and amyloid-PET are PET images, so they share the same image processing step in which we use SPM8 (http://www.fil.ion.ucl.ac.uk/spm/) to spatially normalize each patient’s PET images into the common Montreal Neurological Institute (MNI) altas space. Then, we extract features from each type of PET image separately. From FDG-PET, the features include hypometabolic convergence index (HCI) (Chen et al. 2011), statistical region of interest (sROI) (Chen et al. 2010), and regional precuneus metabolism and posterior cingulate metabolism. All these features have been previously reported to be related to AD (Bailly et al. 2015; Del Sole et al. 2008). From amyloid-PET, the features include SUVRs from six brain regions including orbital frontal cingulate, temporal cortex, anterior cingulate, posterior cingulate, parietal lobe, and precuneus. These regions are known to be associated with amyloid depositions and AD (Fleisher et al. 2011). Because the six SUVRs are highly correlated, we apply principal component analysis (PCA) and include the first PC as a feature for amyloid-PET. Note that IMTL assumes normal distributions of the features. In this application, this assumption naturally holds because each feature is an average of voxel-wise measurements within a brain region. Since many voxels are involved in generating the average, the Central Limit Theorem applies. Also, we generate normal Q-Q plots for the features and find that the normality assumption holds.

#### 4) Inclusion of clinical variables and feature screening

We also include the following clinical variables which could potentially help the early diagnosis and prognosis of AD: age, gender, years of education, APOE e4 status, and cognitive test scores from several commonly used instruments such as mini-mental state examination (MMSE), AD assessment scale-cognitive (ADAS-Cog), clinical dementia rating (CDR), and auditory verbal learning test (AVLT). No patient has missing data for these clinical variables so they are used in the same way as MRI features in our model. Furthermore, we put all the features through a feature screening module using the approach by (Fan and Lv 2008). Note that feature screening is only applied to the training set not the entire dataset to avoid overfitting.

#### 5) Application of IMTL

Within each sub-cohort, the samples are divided into five folds. We combine four folds from each sub-cohort into a training set and use the remaining data as the test set. We apply IMTL to the training set and then use the trained model to predict on the test set. We exhaust all four-fold combinations in training, which produces a 5-fold cross validation procedure for evaluating the accuracy of IMTL. This process is repeated for 50 times. For comparison, two competing methods are applied on the same data: SA and AADM. Table 3 summarizes the results. IMTL has significantly higher AUC and sensitivity than both competing methods in both diagnosis and prognosis. Notably, competing methods have low AUC and sensitivity in prognosis. This is greatly improved by IMTL. Prognosis is more challenging than diagnosis because the former has a heavily imbalanced dataset (46 converters vs. 168 non-converters). Clearly, IMTL is more robust to sample imbalance. All models achieve a similar level of specificity. Finally, we show the contribution of each imaging feature to diagnosis and prognosis by plotting the percentage of times a feature is included in the IMTL model. The result is shown in Fig. 3. Hippocampal volume from MRI and the first PC of six SUVRs from amyloid-PET are almost always included in both diagnostic and prognostic models. This is consistent with findings in the literature that hippocampal atrophy and amyloid-PET SUVRs provide most important biomarkers for AD (Fleisher et al. 2011; Devanand et al. 2007). Other features that are selected for over 50% of the time include HCI, sROI and precuneus metabolism from FDG-PET for diagnosis; and ventricle volume from MRI and HCI and sROI from FDG-PET for prognosis. Clinical variables such as age, APOE e4 status, MMSE, and CDR are more frequently selected. These variables have been widely reported to be related to AD.

**Table 3.** Diagnostic and prognostic performance: ave (std) and p value for hypothesis testing that IMTL is better than a competing method

|  | DIAGNOSIS |  |  | PROGNOSIS |  |  |
| --- | --- | --- | --- | --- | --- | --- |
|  | IMTL | SM | AADM | IMTL | SM | AADM |
| **AUC** | **0.93**(0.03) | 0.86(0.06)<br>p<0.001 | 0.90(0.04)<br>p<0.001 | **0.85**(0.05) | 0.72(0.09)<br>p<0.001 | 0.78(0.07)<br>p<0.001 |
| **SENSITIVITY** | **0.91**(0.06) | 0.84(0.09)<br>p<0.001 | 0.88(0.06)<br>P=0.03 | **0.96**(0.09) | 0.58(0.18)<br>p<0.001 | 0.76(0.17)<br>p<0.001 |
| **SPECIFICITY** | 0.87(0.06) | 0.82(0.06)<br>p<0.001 | 0.86(0.05)<br>P=0.27 | 0.85(0.05) | 0.86(0.05)<br>p=0.78 | 0.86(0.05)<br>P=0.34 |

**Figure 3.**
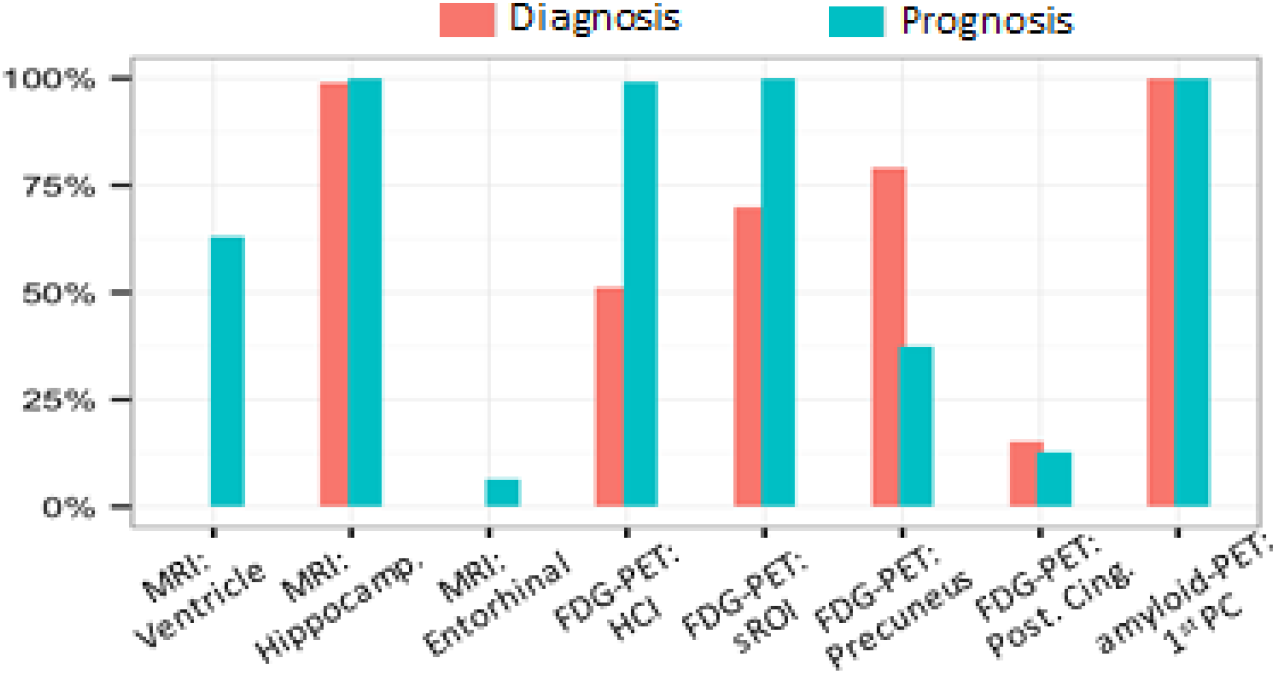
Percentage of times imaging features are included in IMTL over 5 fold cross-validation and 50 repeated experiments.

## 6. Conclusion

In this paper, we proposed IMTL to build predictive and classification models for IMD data. We developed an EM algorithm for parameter estimation of IMTL and further extended it to achieve between-institutional collaborative model estimation without the need for data pooling. We demonstrated that IMTL was capable of out-of-sample prediction and proved that it had a larger Fisher information than models without transfer learning under mild conditions. This explained the superiority of IMTL from a theoretical standpoint. Simulation experiments demonstrated high accuracy in using IMTL for out-of-sample prediction and classification. IMTL was applied to AD early diagnosis and prognosis, i.e., at the MCI stage, using incomplete multimodality imaging data. Significantly higher AUC and sensitivity were achieved in both diagnosis and prognosis compared with competing methods. Image features selected to include in the models were widely reported in the literature to be related to AD.

This research has several limitations: First, IMTL assumes normal distributions of features. To make IMTL an appropriate choice for an application, feature normality needs to be checked. If the features do not follow a normal distribution, cox transformation may be used. Nevertheless, extension of IMTL to non-normal features provides a more general approach, and thus being an interesting future research direction. Second, this paper focuses on response variables that are either normal or binary. Extending the current modeling framework to other types of response variables will be valuable to address the need of various application domains. Third, IMTL assumes equal variance-covariance for the models in different sub-cohorts. This assumption can be relaxed to accommodate potential sub-cohort heterogeneity.

## Data Availability

Data used in the preparation of this article were obtained from the Alzheimer’s Disease Neuroimaging Initiative (ADNI) database (adni.loni.usc.edu). As such, the investigators within the ADNI contributed to the design and implementation of ADNI and/or provided data but did not participate in analysis or writing of this report. A complete listing of ADNI investigators can be found at: http://adni.loni.usc.edu/wp-content/uploads/how_to_apply/ADNI_Acknowledgement_List.pdf

## Appendix

### A. Proof of Theorem 1

Under IMTL, the complete-data log-likelihood function is:

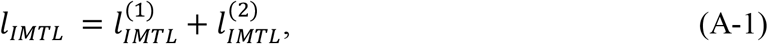

where 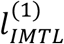 and 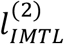 correspond to the two sub-cohorts, i.e.,

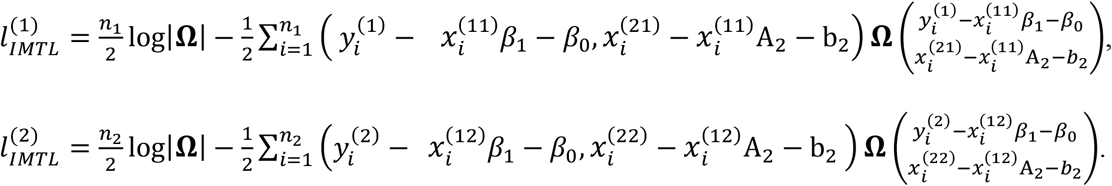

Because (A-1) includes missing data, computing the observed Fisher information needs to take the expectation of D^*mis*^ given D^*obs*^ and the parameters. This follows from the discussion in Chapter 7 of the book by Little and Rubin (2002) on Fisher information with missing data. Specifically, the observed Fisher Information for each element *Ω*_*ij*_ in **Ω** is

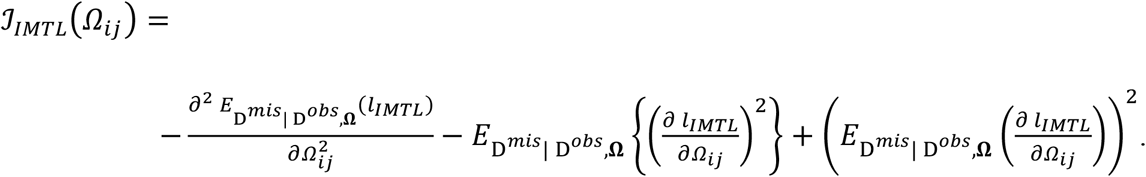

Applying this formula to *Ω*_22_, *Ω*_2*y*_, and *Ω*_*yy*_, respectively, we can get

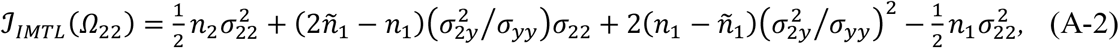

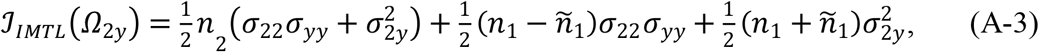

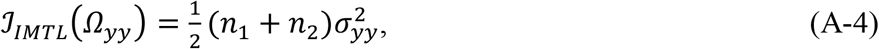

where

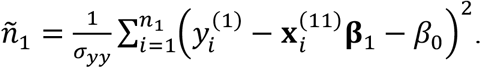

Under SM, the model for sub-cohort 2 and the corresponding log-likelihood function are the same as those by IMTL, i.e.,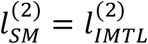. However, sub-cohort 1 is modeled separately from sub-cohort 2, as shown in (22), with corresponding log-likelihood function being

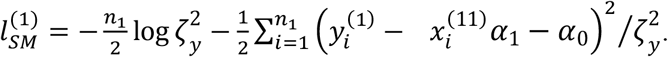

Taking the two sub-cohorts together, the log-likelihood function of SM is:

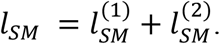

Since *l*_*SM*_ does not include any missing data, the observed Fisher information can be computed in the regular way (Little and Rubin (2002)), i.e.,

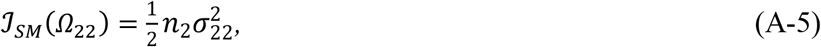

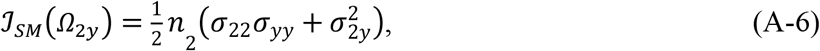

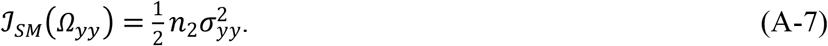

Under AADM, the model for sub-cohort 1 has the same form as SM but with a different log-likelihood function due to the incorporation of the data from sub-cohort 2, i.e.,

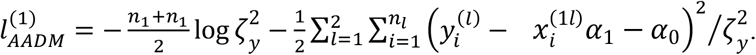

Furthermore, due to the same reasons as SM, 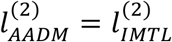. Taking the two sub-cohorts together, the log-likelihood function of AADM is:

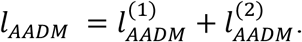

It is not hard to recognize that the observed Fisher information of AADM has the same formula as SM in (A-5), (A-6), and (A-7). This is because computing the observed Fisher information of *Ω*_22_, *Ω*_2*y*_, and *Ω*_*yy*_ only concerns the log-likelihood function of sub-cohort 2, which is the same for SM and AADM.

Furthermore, we take the expectation of the observed Fisher information in each model with respect to *D*_*obs*_, which produces the Fisher information. Comparing the Fisher information between IMTL and SM (or AADM), we have

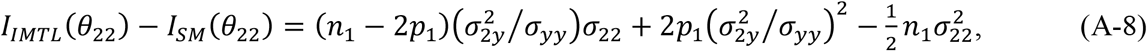

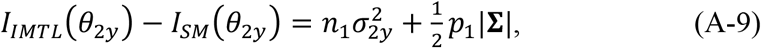

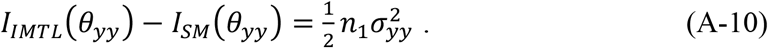

The right-hand sides of (A-9) and (A-10) are positive, which means *I*_*IMTL*_(*θ*_2*y*_) > *I*_*SM*_(*θ*_2*y*_) and *I*_*IMTL*_(*θ*_*yy*_) > *I*_*SM*_(*θ*_*yy*_) unconditionally. To make *I*_*IMTL*_(*θ*_22_) > *I*_*SM*_(*θ*_22_), the right-hand side of (A-8) needs to be positive, which means

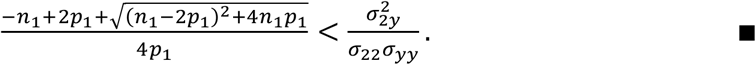

### B. IMTL classification model: formula for EM estimation and classification on new samples

Here, we skip the details and present the result of derivations in the E-step and M-step. Specifically, the E-step derives the following expectations:

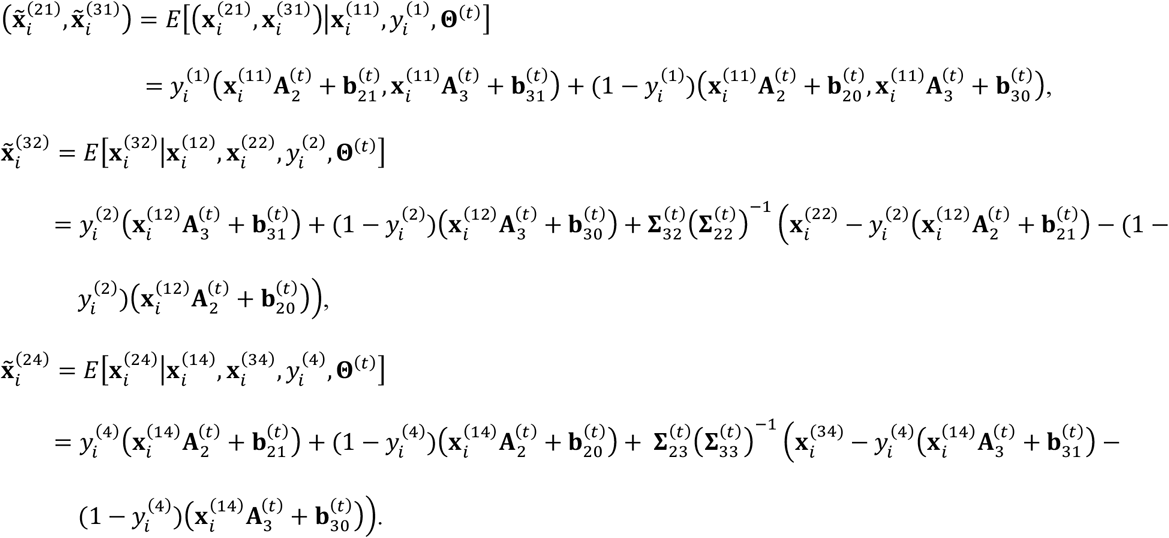

In the M-step, the parameters in **Θ** can be updated as

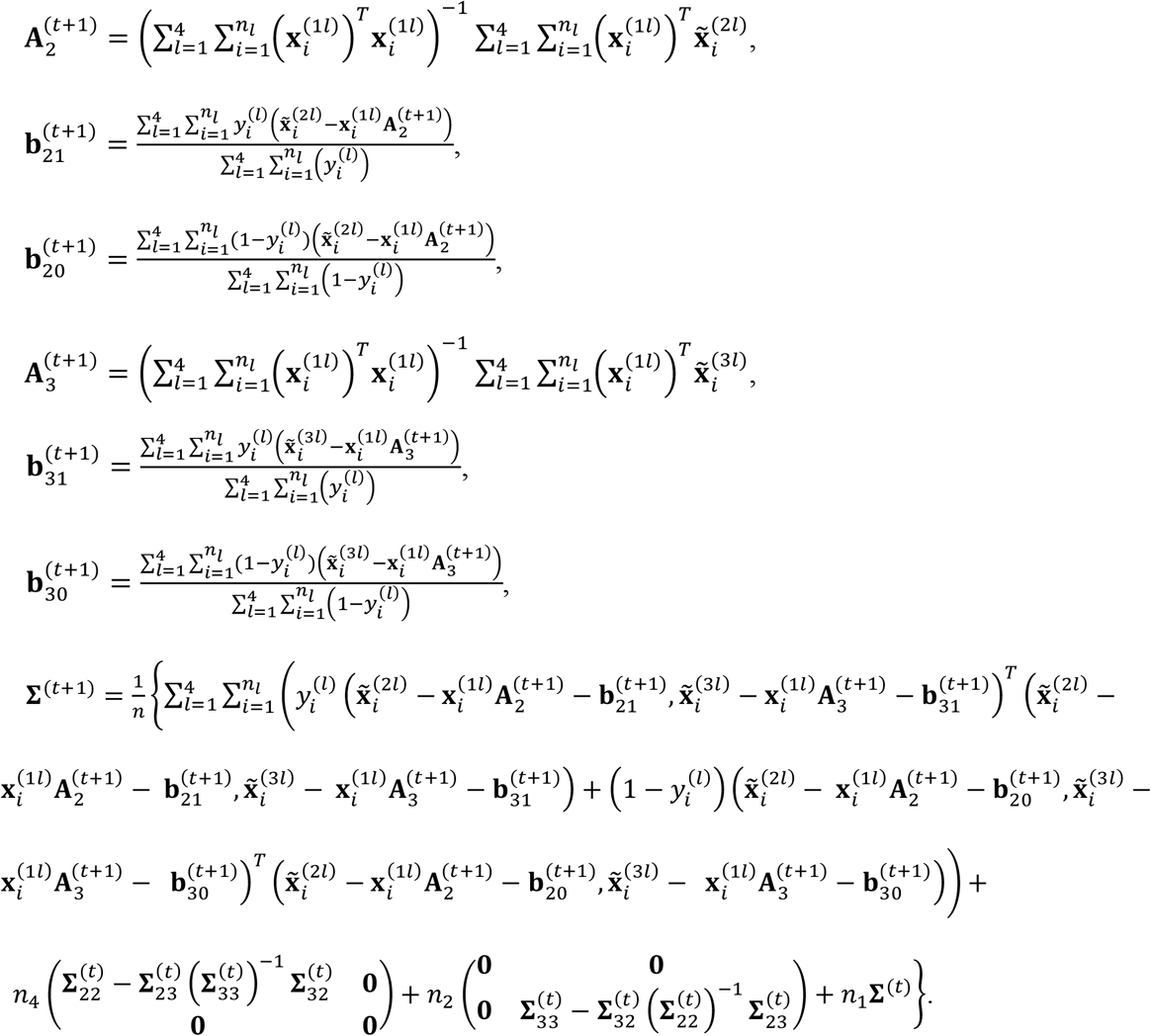

At the convergence of the above EM iterations, we can obtain the estimated 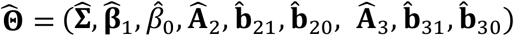. These parameters can be used in a logistic regression model to predict on new samples. Consider a new sample *i*^∗^. Depending on what available modality/modalities this sample has, i.e., which sub-cohort the sample belongs to, we can use the following model to predict the response variable of the sample:

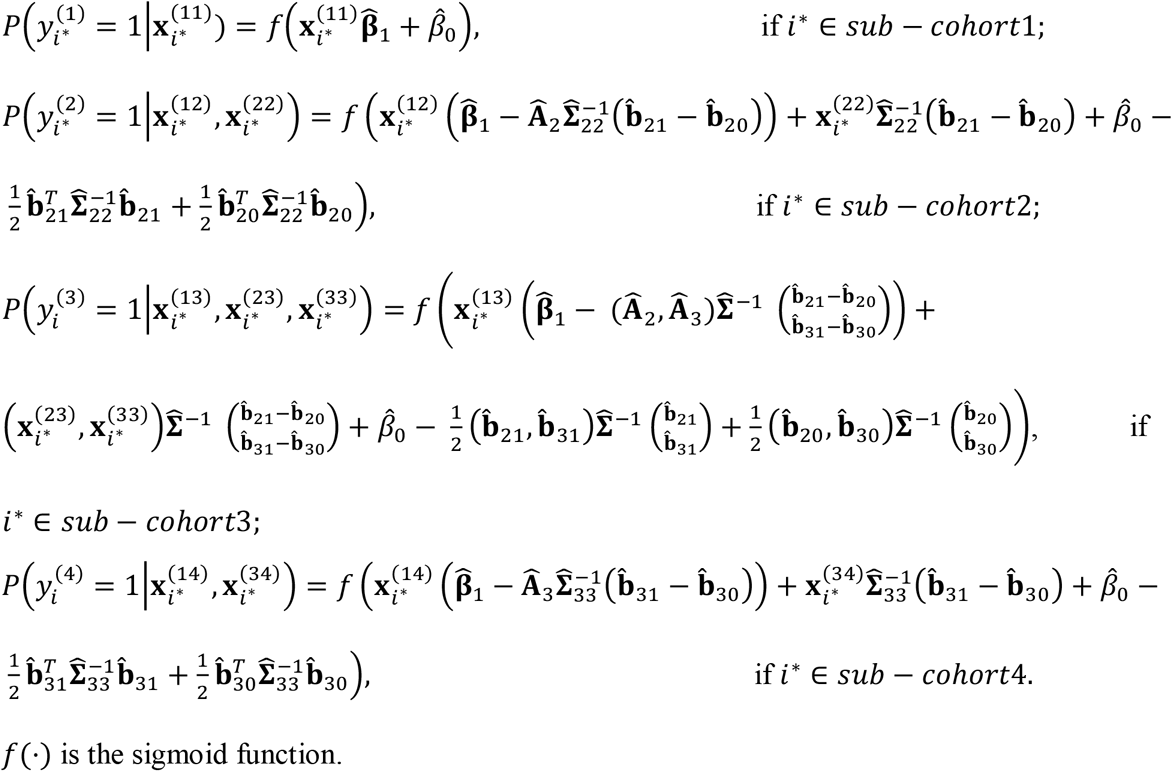

## Acknowledgements

This research is partly supported by NIH-NIA grant no. R41AG053149 and NSF CAREER 1149602. Data collection and sharing for this project was funded by the Alzheimer’s Disease Neuroimaging Initiative (ADNI) (National Institutes of Health Grant U01 AG024904) and DOD ADNI (Department of Defense award number W81XWH-12-2-0012). ADNI is funded by the National Institute on Aging, the National Institute of Biomedical Imaging and Bioengineering, and through generous contributions from the following: AbbVie, Alzheimer’s Association; Alzheimer’s Drug Discovery Foundation; Araclon Biotech; BioClinica, Inc.; Biogen; Bristol-Myers Squibb Company; CereSpir, Inc.; Cogstate; Eisai Inc.; Elan Pharmaceuticals, Inc.; Eli Lilly and Company; EuroImmun; F. Hoffmann-La Roche Ltd and its affiliated company Genentech, Inc.; Fujirebio; GE Healthcare; IXICO Ltd.; Janssen Alzheimer Immunotherapy Research & Development, LLC.; Johnson & Johnson Pharmaceutical Research & Development LLC.; Lumosity; Lundbeck; Merck & Co., Inc.; Meso Scale Diagnostics, LLC.; NeuroRx Research; Neurotrack Technologies; Novartis Pharmaceuticals Corporation; Pfizer Inc.; Piramal Imaging; Servier; Takeda Pharmaceutical Company; and Transition Therapeutics. The Canadian Institutes of Health Research is providing funds to support ADNI clinical sites in Canada. Private sector contributions are facilitated by the Foundation for the National Institutes of Health (www.fnih.org). The grantee organization is the Northern California Institute for Research and Education, and the study is coordinated by the Alzheimer’s Therapeutic Research Institute at the University of Southern California. ADNI data are disseminated by the Laboratory for Neuro Imaging at the University of Southern California.

